# The incubation period of 2019-nCoV infections among travellers from Wuhan, China

**DOI:** 10.1101/2020.01.27.20018986

**Authors:** Jantien A. Backer, Don Klinkenberg, Jacco Wallinga

## Abstract

Currently, a novel coronavirus 2019-nCoV causes an outbreak of viral pneumonia in Wuhan, China. Little is known about its epidemiological characteristics. Using the travel history and symptom onset of 88 confirmed cases that were detected outside Wuhan, we estimate the mean incubation period to be 6.4 (5.6 – 7.7, 95% CI) days, ranging from 2.1 to 11.1 days (2.5^th^ to 97.5^th^ percentile). These values help to inform case definitions for 2019-nCoV and appropriate durations for quarantine.

---

Early January 2020, a novel coronavirus 2019-nCoV, has been identified as the infectious agent that causes an outbreak of viral pneumonia in Wuhan, China, where the first cases had their symptom onset in December 2019 [1]. This newly discovered virus causes a severe acute respiratory disease. The rapid spread in Wuhan and throughout China poses a great strain on health care system and disrupts the local societies. The multiple cases showing up outside China cause international concern.

The virus is related to the SARS coronavirus and MERS coronavirus, but is distinct from each of those viruses [2], such that the epidemiological key parameters have to be identified for this new virus from incoming case reports as the epidemic continues. Chief among these key parameters is the incubation period distribution. The range of the values for the incubation period is essential to epidemiological case definitions, and is required to determine the appropriate duration of quarantine. Moreover, knowledge of the incubation period helps to assess the effectiveness of entry screening and contact tracing. The distribution of the incubation period is also used in estimating the size of the epidemic [3 – 5] and the transmission potential [6, 7]. In absence of data on the 2019-nCoV incubation period, the WHO has worked with a broad range of 0 to 14 days, the CDC has taken a range of 2 to 14 days, and several studies have assumed incubation periods of SARS or MERS coronaviruses.

Here we present the distribution of incubation periods estimated for travellers from Wuhan with confirmed 2019-nCoV infection, using their reported travel histories and symptom onset dates.

## Travellers from Wuhan with confirmed 2019-nCoV infection, a reported symptom onset data and a reported travel history

In January 2020 an increasing number of cases were detected outside Wuhan, China and confirmed to be infected with 2019-nCoV. For 88 cases detected between 20 and 28 January, the travel history (to and) from Wuhan is known as well as their symptom onset date. Their ages range from 2 to 72 years old (with four missing), 31 of them are women, 57 are men. During this initial stage of the epidemic, it is most likely that these travellers were infected in Wuhan. Consequently, their time spent in Wuhan can be taken as the duration of exposure to infection.

Of these 88 cases with known travel history, 63 were Wuhan residents and travelled elsewhere, and 25 visited Wuhan for a limited time. By taking the date of symptom onset and travel history together we inferred the possible values that the incubation period could have been for each of these cases. Examples of these reports are:

“*First confirmed imported nCov pneumonia patient in Shanxi: male, visited Wuhan from 01/12/2020 to 01/15/2020, symptom onset on 01/19/2020, visit clinic on 01/20/2020, 6 contacts traced*.*”* which gives an observed incubation period between 4 and 7 days.

*“First confirmed imported nCov pneumonia patient in Shanghai (from Wuhan): female, 56, Wuhan residence, arrived in Shanghai from Wuhan on 01/12/2020, symptom onset and visited fever clinic on 01/15/2020, laboratory confirmed on 01/20/2020”* which gives an observed incubation period of 3 days or longer.

The data used for this analysis has been translated from Chinese sources and made publicly available [8]. We took the data as available on January 29, 2020 (supplementary material S1).

## Incubation period distribution

Using the duration of stay in Wuhan and the symptom onset date, we obtained a range of possible values for the incubation period of each case. We fitted three parametric forms for the incubation period distribution to these ranges: the Weibull distribution, the gamma distribution, and the lognormal distribution. We used a Bayesian approach to fitting, that allows for the use of prior knowledge to inform the analysis. We specified strictly positive flat prior probability distributions for the parameter values of the three distributions (supplementary material S2), which ensured our estimates are conservative. Because of the high number of observations, the impact of the priors on the outcome was negligible. We used a uniform prior probability distribution over the exposure interval for the moment of infection for each case. We sampled from the posterior distribution using the rstan package [9] in R [10] (supplementary material S3).

Figure 1 shows the time line for each case, where the cases without a maximum incubation period lack an unexposed (gray) period. However, the estimated infection times for these cases are close to the end of the exposure window, informed by the cases that do have a maximum incubation period.

**Fig. 1.**
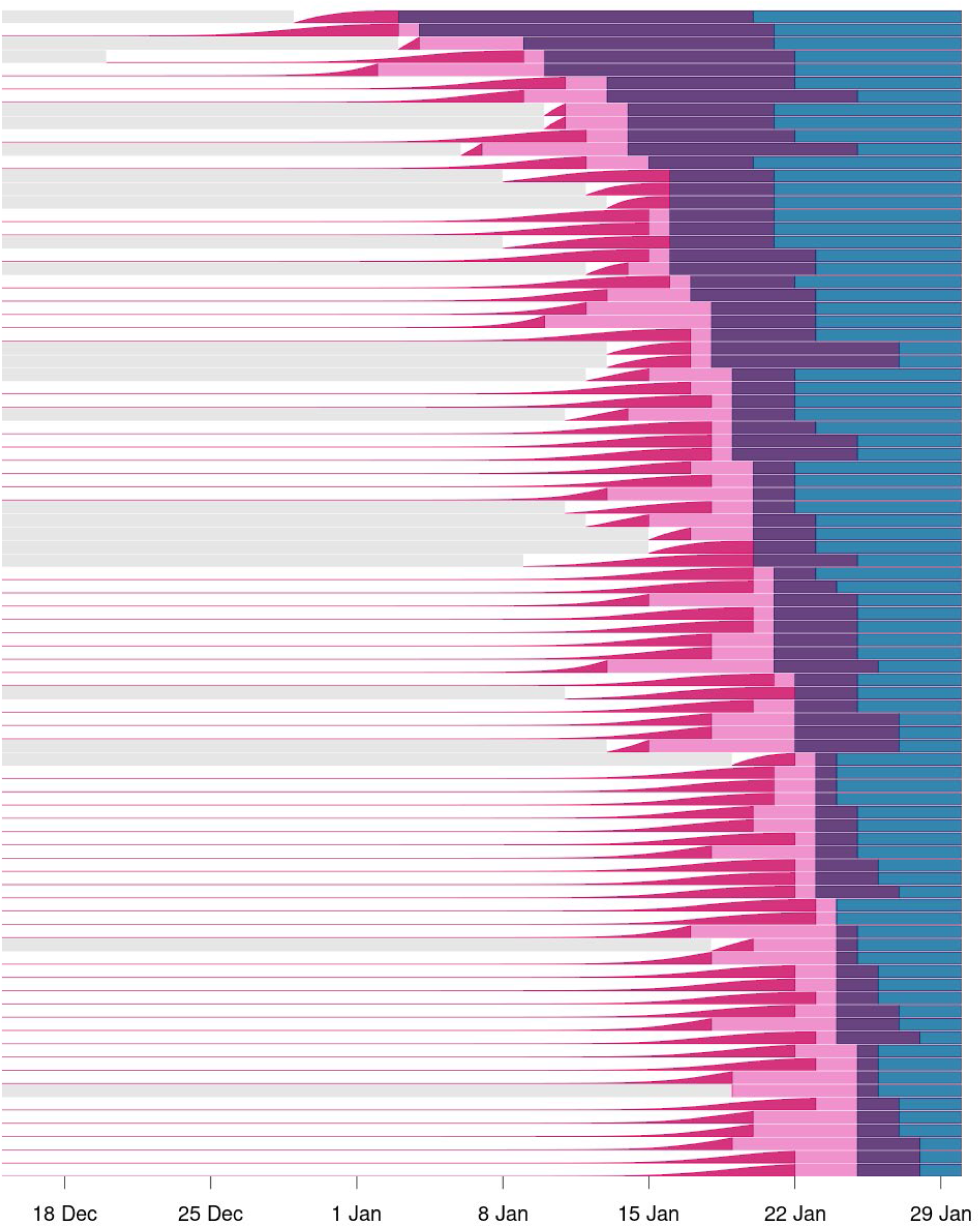
Time line for each case with travel history from Wuhan, sorted by symptom onset date. Distinct periods are shown as unexposed (gray), exposed (white), infected but asymptomatic (pink), symptomatic (purple) and reported (blue). The analysis yields the probability of being infected (violet), i.e. the cumulative density function of the estimated infection moments (using the Weibull distribution).

The Weibull distribution provided the best fit to the data (Tab. 1). The mean incubation period was estimated to be 6.4 (5.6 – 7.7, 95% credible interval) days. The incubation period ranges from 2.1 to 11.1 days (2.5^th^ to 97.5^th^ percentile) (Tab. 2 and Fig. 2). The results using the gamma distribution provide a slightly poorer description of the data but provide a similar range from 2.4 to 12.5 days. Although the lognormal distribution provides the poorest fit to the data, the incubation period ranging from 2.4 to 15.5 days (2.5^th^ to 97.5^th^ percentile) may be relevant for a conservative choice of quarantine periods.

**Tab. 1.** Estimated incubation period for travellers infected with 2019-nCoV in Wuhan, China, January 2020, for different parametric forms for the likelihood; posterior median and 95% credible interval in days.

| Distribution | mean (days) |  | sd (days) |  | LOO IC* |
| --- | --- | --- | --- | --- | --- |
|  | estimate | (95% CI) | estimate | (95% CI) |  |
| Weibull | 6.4 | (5.6 – 7.7) | 2.3 | (1.7 – 3.7) | 486 |
| Gamma | 6.5 | (5.6 – 7.9) | 2.6 | (1.8 – 4.2) | 545 |
| Lognormal | 6.8 | (5.7 – 8.8) | 3.4 | (2.1 – 6.4) | 592 |

\* LOO IC: Leave-One-Out Information Criterion; this indicates the goodness-of-fit, where lower values indicate a better fit and differences larger than two are statistically relevant.

**Tab. 2.** Percentiles of estimated incubation period for travellers infected with 2019-nCoV in Wuhan, China, January 2020, for different parametric forms for the likelihood; posterior median and 95% credible interval in days.

| Percentiles | Weibull |  | Gamma |  | Lognormal |  |
| --- | --- | --- | --- | --- | --- | --- |
|  | estimate | (95% CI) | estimate | (95% CI) | estimate | (95% CI) |
| 2.5<sup>th</sup> | 2.1 | (1.3 – 3.0) | 2.4 | (1.5 – 3.2) | 2.4 | (1.6 – 3.1) |
| 5<sup>th</sup> | 2.7 | (1.8 – 3.5) | 2.9 | (2.0 – 3.6) | 2.8 | (2.0 – 3.5) |
| 50<sup>th</sup> | 6.4 | (5.5 – 7.5) | 6.1 | (5.3 – 7.3) | 6.1 | (5.2 – 7.4) |
| 95<sup>th</sup> | 10.3 | (8.6 – 14.1) | 11.3 | (9.1 – 15.7) | 13.3 | (9.9 – 20.5) |
| 97.5<sup>th</sup> | 11.1 | (9.1 – 15.5) | 12.5 | (9.9 – 17.9) | 15.5 | (11.0 – 25.2) |
| 99<sup>th</sup> | 11.9 | (9.7 – 17.2) | 14.1 | (10.9 – 20.6) | 18.5 | (12.6 – 32.2) |

**Fig. 2.**
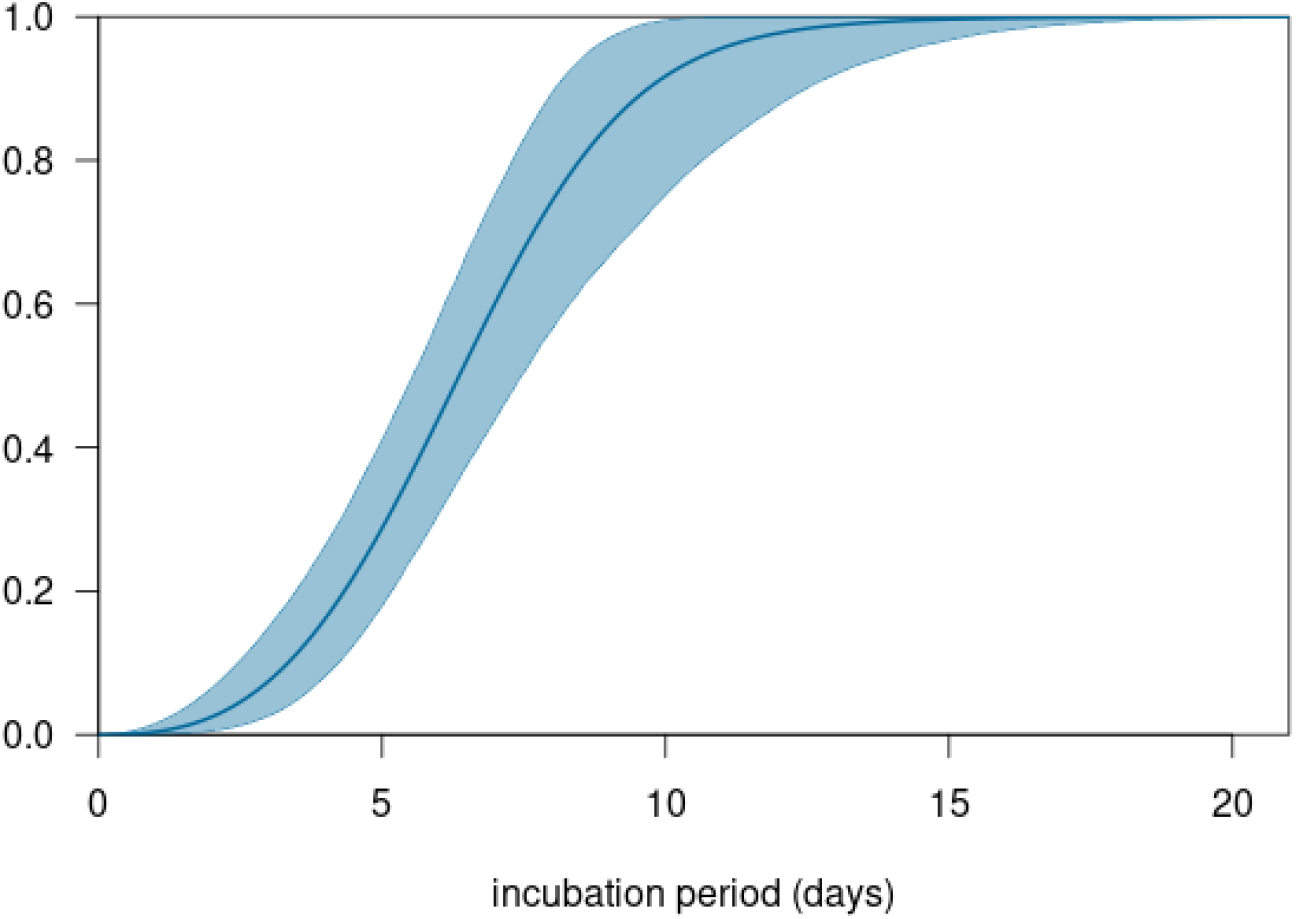
The cumulative density function of the estimated Weibull incubation period distribution for travellers infected with the 2019-nCoV in Wuhan, China, January 2020; posterior median of mean (blue line) and 95% credible interval (light blue area).

## Comparison of 2019-nCoV incubation period to incubation periods of SARS and MERS coronaviruses

A comparison to the estimated incubation period distribution for MERS (Tab. 3 and Fig. 3) shows that the values are remarkably similar, with mean values differing at most 1 day and 95^th^ percentiles differing at most 2 days. The estimated mean incubation periods for SARS are more variable between studies, including values shorter and longer than presented here for 2019-nCoV. These findings imply that findings of previous studies that have assumed incubation period distribution similar to MERS or SARS will not have to be adapted because of a shorter or longer incubation period.

**Tab. 3.** Estimated incubation periods for coronaviruses from different studies

| study | Virus<br>(location) | Distribution | mean (days) |  | 95<sup>th</sup> percentile (days) |  |
| --- | --- | --- | --- | --- | --- | --- |
|  |  |  | estimate | (95% CI) | estimate | (95% CI) |
| this study | nCoV | Weibull | 6.4 | (5.6 – 7.7) | 10.3 | (8.6 – 14.1) |
| this study | nCoV | Gamma | 6.5 | (5.6 – 7.9) | 11.3 | (9.1 – 15.7) |
| this study | nCoV | Lognormal | 6.8 | (5.7 – 8.8) | 13.3 | (9.9 – 20.5) |
| Donnelly 2003<br>[11] | SARS | Gamma | 6.4 | (5.2 – 7.7) | 14.2 | n.a. |
| Cowling 2007<br>[12] | SARS | Lognormal | 5.1 | (4.6 – 5.5) | 12.9 | (11.7 – 14.5) |
| Lau 2010 [13] | SARS<br>(Hong Kong) | Lognormal | 4.4 | n.a. | 12.4 | n.a. |
| Lau 2010 [13] | SARS (Beijing) | Lognormal | 5.7 | n.a. | 19.7 | n.a. |
| Lau 2010 [13] | SARS (Taiwan) | Lognormal | 6.9 | n.a. | 17.9 | n.a. |
| Lessler 2009 [14] | SARS | Lognormal | 4.8<sup>#</sup> | (3.6 – 4.4) | 10.6 | (8.9 – 12.2) |
| Assiri 2013 [15] | MERS | Lognormal | 6.0<sup>†</sup> | (1.9 – 14.7) | 12.4 | (7.3 – 17.5) |
| Cauchemez<br>2014 [16] | MERS | Lognormal | 5.5 | (3.6 – 10.2) | 10.2<sup>*</sup> | n.a. |
| Virlogeux 2016<br>[17] | MERS<br>(South Korea) | Gamma | 6.9 | (6.3 – 7.5) | 12.7 | (11.5 – 14.4) |
| Virlogeux 2016<br>[17] | MERS<br>(Saudi Arabia) | Lognormal | 5.0 | (4.0 – 6.6) | 11.4 | (8.5 – 17.5) |

<sup>#</sup> Value calculated from median of 4.0 (3.6 – 4.4) days provided in reference
<sup>†</sup> Value calculated from median of 5.2 (1.9 – 14.7) days provided in reference
<sup>\*</sup> Value calculated from mean and sd provided in reference

**Fig. 3.**
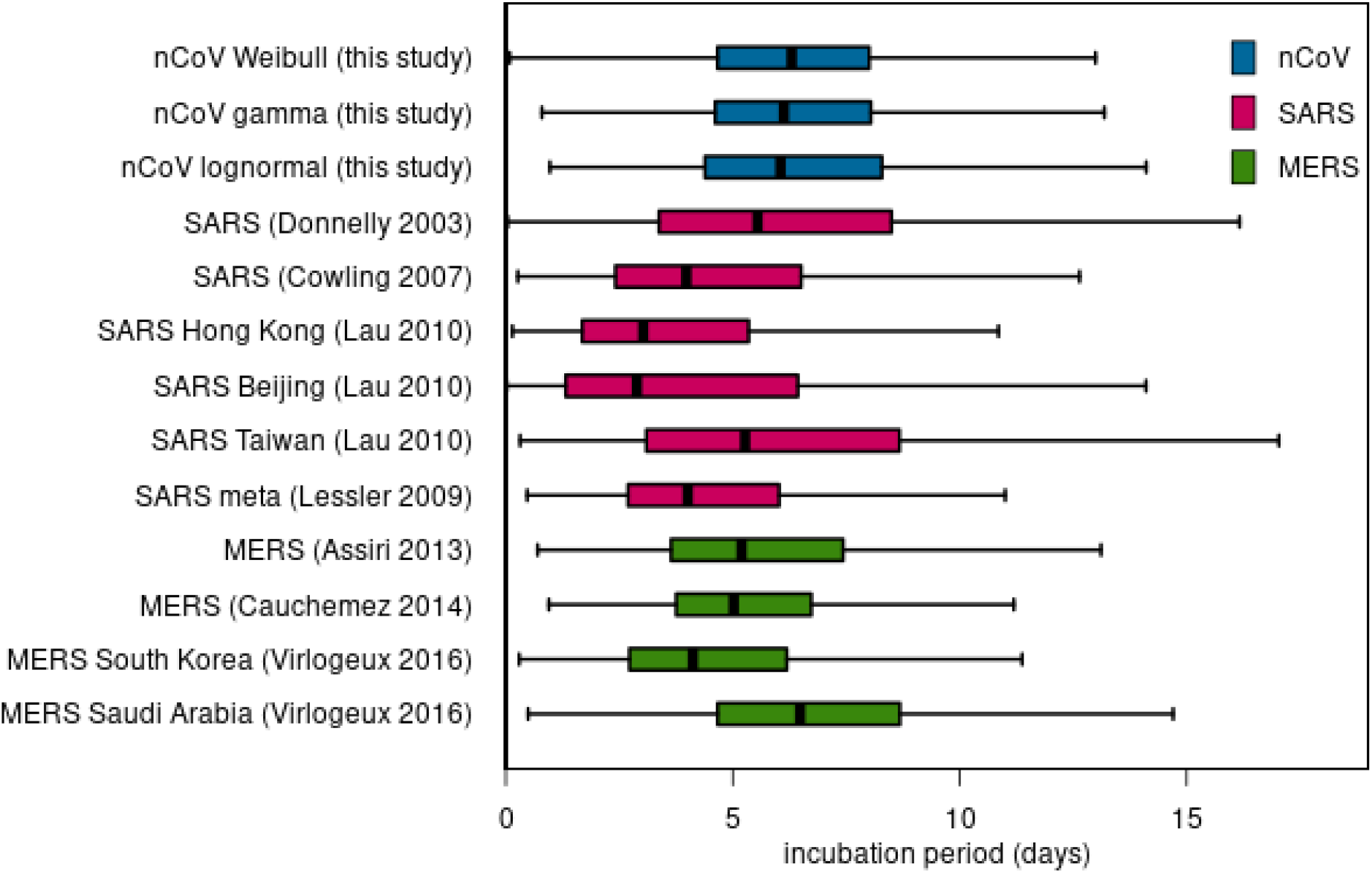
Box-and-whisker-plots of estimated incubation periods for coronaviruses from different studies; median (black point), interquartile range (box), and maximum of 1.5 times the interquartile range (whiskers).

## Discussion

We characterized the distribution of incubation periods for Chinese travellers infected with 2019-nCoV in Wuhan, who were reported as cases between 20 and 28 January 2020. The study provides empirical evidence to back reports on a familial cluster where five family members developed symptoms 3 – 6 days after exposure [18] and a reported range from 2 to 11 days based on 16 travellers between Wuhan and Guangdong [19]. It fits within the range from 0 to 14 days that has been assumed by the WHO.

In our analysis we assumed a uniform prior probability of being infected during the period of stay in Wuhan. Since the epidemic was developing during that time period, it is more likely that travellers were infected toward the end rather than the beginning of their stay. This might produce a slight bias towards longer incubation periods, so the estimated upper limit of 11.1 days could be considered conservative.

The travellers in this study represent a selective sample of the reported cases. We found travellers to be more often male and younger than the cases reported. The numbers are too small to detect systematic differences in incubation time with age or sex. Because we only have information on confirmed cases, there is likely a bias towards more severe cases in areas with a functioning health care system. As the epidemic continues, it remains important to collect more information on incubation periods of 2019-nCoV cases of older ages, of persons with underlying morbidity, of women, and of persons with mild symptoms.

There are various choices one can make about the parametric form of the incubation period distribution. The results with the three often-used forms we report here suggest that there is little impact on the mean and dispersion of the incubation periods. Of these three, the lognormal distribution assigns higher probabilities to longer incubation periods. Although we found that this distribution provided a poorer description of the data than the Weibull and the gamma distributions, it is prudent not to dismiss the possibility of incubation periods up to 14 days at this stage of the epidemic.

Our study helps in choosing an appropriate duration for quarantine. Keeping infected persons in quarantine up to 14 days will ensure that they do not develop symptoms after release.

## Data Availability

The data used for this analysis has been translated and made publicly available by Dr. Kaiyuan Sun (NIH, USA). We took the data as available on January 29 (supplementary material S1).

https://docs.google.com/spreadsheets/d/1jS24DjSPVWa4iuxuD4OAXrE3QeI8c9BC1hSlqr-NMiU/edit#gid=1449891965

## Acknowledgments

We are immensely indebted to the work of Dr. Kaiyuan Sun, Ms. Jenny Chen, Dr. Cécile Viboud and the staff of MOBS Lab led by Prof. Alessandro Vespignani who traced and translated the information and generously made it available to the wider community.

